# Effect of Performance-Based Financing on the quality of Obstetric Service delivery in Nkambe Health District, North Region, Cameroon

**DOI:** 10.64898/2026.02.27.26347217

**Authors:** Eneigheo Emmanuel Achuondou, Obase Ralph, Nji Valery Che, Bodzewan Emmanuel Fonyuy

## Abstract

**Background:** Performance-based financing (PBF) aims to improve healthcare quality by offering financial incentives to service providers. This study evaluated PBF’s impact on obstetric care quality in Nkambe Health District, Cameroon.

**Methods:** We employed a cross-sectional mixed-methods study from December 2016–July 2017 in PBF-implementing facilities. We surveyed 315 women attending prenatal and postnatal visits and 23 service providers using structured questionnaires. Quantitative data were analyzed with SPSS v20, and quality was assessed via WHO standards.

**Results:** 99.7% of women attended ≥5 antenatal visits; 93% delivered institutionally; 93% had postnatal care. Partogram use was 88%; 96% women reported satisfaction. Providers’ knowledge was high (97% for ANC/intrapartum care). Schooling (p<0.05), ≥4 ANC visits (p=0.017), and proximity predicted quality. Providers (71%) were satisfied with pay.

**Conclusions:** PBF significantly enhanced obstetric care utilization and quality. Scale-up recommended with attention to costs, staffing, and attitudes.

## 1 Introduction

According to the United Nations International Children’s Emergency Fund (UNICEF),there were 6.6 million under five deaths in 2012 globally, a remarkable decline from the 12.6 million deaths recorded in 1990(1).Despite the impressive improvements in most regions, the progress made was not sufficient to meet the MDG’s targets due to variations in the rate of reduction in various regions. For instance, although Africa showed encouraging progress in reducing child mortality, 3.2 million deaths were recorded in 2012, accounting for almost half of all child deaths globally, an increase of 29 %(2)

Cameroon, signed the United Nations Millinium Development goals in September 2000, comprising of goals and targets and indicators to be achieved by 2015 using 1990 as a reference. Health related goals were to reduce under 5 mortality by two-thirds, reduce maternal mortality ratio by three quarters and improved universal access to reproductive health(3,3)

In Cameroon, the situation was not different, while sub-Saharan Africa’s annual under-five mortality rate reduction was over five times faster between 2005 and 2013 than it was during 1990–1995(4); Cameroon witnessed an increase of under 5 mortality rate from 772 in 2004 to 782 deaths per 100,000 live births in 2011(2).Similarly, infant mortality rates was 103 per 1000 live births against a national target of 38 per 1000(5).Birth asphyxia,prematurity,sepsis, and congenital anomalies were the principal causes of infant mortality(5). Moreso, 51% of all under-five deaths occurred before the child’s first birthday(2).Studies showed that under-five mortality rates were twice as high for children in the poorest households compared to those from the richest households. In addition, children in rural areas were 1.7 times more likely to die before their fifth birthday than those in urban areas. Children from mothers with secondary or higher education were almost three times as likely to survive than children from mothers with no education.(2)

Many African countries have demonstrated either stagnating or slow progress in neonatal mortality rates. Curbing neonatal mortality is critical to improve the survival of children. Among the proven, cost-effective, and high-impact interventions are skilled care at birth and emergency obstetric care, management of preterm births, basic neonatal care, neonatal resuscitation, early antibiotic treatment for serious infections, inpatient care for small and sick newborns, and the prevention of mother-to-child transmission) of HIV(4).Combining quality health services at the health facility, supported by strong outreach, follow-up, and referral services, promoting healthy behaviors at home, and making early decisions to seek care will have the greatest impact.(1)

Maternal survival has significantly improved since the adoption of MDGs. The maternal mortality ratio decreased from 380 maternal deaths per 100,000 live births to 210, a 45% decrease Worldwide between 1990 and 2013(2).Many developing regions with the highest maternal mortality ratios have also made steady progress in improving maternal health. For example, in sub-Saharan Africa, the maternal mortality ratio declined by 49% between 1990 and 2013, same trend with Southern Asia, which fell by 64%.

The World Health Organization (WHO) reported 800 maternal deaths per 100,000 live births in 2013 alone due to complications of pregnancy and childbirth. It was indicated that the risk of a woman in a developing country, especially in Africa, dying from a maternal cause was approximately 23 times higher than that of a woman in a developed country. Maternal mortality in Africa has been largely associated with three kinds of delays in the childbearing process;delays in seeking healthcare, delays in reaching caregivers, and delays in receiving care (ECA et al., 2011).

Between 1990 and 2013, only four African countries reduced their Maternal Mortality Ratio (MMR) by over 75 %, thereby meeting MDG 5: Cabo Verde, 77 %; Equatorial Guinea, 81.9 %; Eritrea, 77.6 %; and Rwanda, 77.1 %. Some countries reduced their MMR by over 60 % and were on track to meet MDG 5: Ethiopia, 70 %; Angola, 67 %; Mozambique, 63 %; Egypt, 62.5 %; and Morocco, 61.3 %(2)

Globally, there were an estimated 289,000 maternal deaths in 2013, equivalent to approximately 800 deaths each day. Sub-Saharan Africa and Southern Asia, both accounted for 86% of all maternal deaths globally in 2013, most of which were preventable. Hemorrhage accounted for the greatest number of maternal deaths, responsible for more than 27% of maternal deaths in the developing regions and approximately 16% in the developed regions. Other major complications included infections, high blood pressure, complications from delivery, and unsafe abortion(5)

Additionally, more than 71 % of births were assisted by skilled health personnel globally in 2014, an increase of 59 % from 1990. In the developing regions, only 56 % of births in rural areas were attended by skilled health personnel, compared with 87 % in urban areas(2). Similarly, half of the pregnant women in developing regions received the recommended minimum of four antenatal care visits. Moreover, only 51% of countries have data on maternal causes of death. In Cameroon, maternal mortality rose from 778 in 2004 to 782 in 2011, in contrast to global trends and mimicking sub-Saharan Africa(4).Proven healthcare interventions that can prevent or manage these complications, including antenatal care during pregnancy, skilled care during childbirth, and care and support in the weeks after childbirth (6).

In Cameroon, maternal mortality rate increased from 669 per 100,000 live births in 2005 to 1000 per 100,000 live births in 2010(7).Research evidence has identified various factors that contribute to maternal mortality, including socioeconomic, cultural, and obstetric factors. Direct causes such as postpartum hemorrhage, unsafe abortions, and hypertensive disorders in pregnancy have been reported as the leading causes of maternal deaths in many developing countries, while indirect causes such as severe malaria and HIV/AIDS have been found to play a significant role in maternal mortality in sub-Saharan Africa(2)

High maternal and child mortality rates profoundly affect a country’s social, economic, and developmental progress(2).Maternal deaths often lead to financial instability in families, as women frequently contribute significantly to household income. The loss of a mother can result in decreased productivity and increased healthcare costs, thereby hindering economic growth. Investments in maternal and child health have been shown to yield substantial economic benefits; for instance, every dollar spent can generate up to $7 in economic returns owing to improved health outcomes and productivity(8).The death of the mother significantly increases the risk of mortality. Children who lose their mothers are more susceptible to malnutrition, illness, and lack of education, which can perpetuate cycles of poverty and poor health (6)

High maternal and child mortality rates often indicate systemic issues within a country’s healthcare infrastructure, such as inadequate access to quality care, insufficient medical supplies, and shortage of trained healthcare professionals. Addressing these challenges requires substantial investment and reform to improve health care delivery and outcomes (2).Elevated mortality rates can further lead to demographic imbalances and social challenges, including an increase in the number of orphans and disrupted family structures. These outcomes can limit social services and impede community development (9).Maternal mortality can negatively impact children’s education, particularly for girls who may leave school to assume caregiving responsibility. This disruption can hinder efforts toward gender equality and limit opportunities for women’s empowerment (10)

Emergency obstetric care is crucial for reducing maternal and neonatal mortality through targeted interventions during pregnancy, delivery, and the post-natal period. Evidence-based EmOC services manage potentially life-threatening complications that affect many women during pregnancy, childbirth, and the immediate postpartum period.(2)

To accelerate progress toward meeting the MDGs goals, developing countries need to increase access to and quality of maternal and child health services. An intervention that showed promise for improving access to and quality of health services was performance-based financing (PBF)(10). PBF schemes provide financial incentives to healthcare providers for improvements in utilization and quality of specific care indicators, and influence the provision of health care in two ways: first, by giving incentives to service providers to put more effort into maternal and child services, and second, by increasing the amount of resources available to finance the delivery of services.(11)

PBF has attracted considerable interest from government and aid agencies in low-income countries as a means to increase the productivity and quality of healthcare providers. In Africa alone, more than 35 countries, including Cameroon, have implemented PBF(12).In Rwanda, for example, PBF has proven to be an efficient way to increase health service quality and utilization, resulting in improved child health outcomes, while in the Democratic Republic of Congo, Elise Huillery et al. (2013) found that financial incentives improved efforts from health workers to increase targeted service provision, but demand for health services was not responsive to these incentives. Loevinsohn and Harding (2005) reviewed studies on the effect of contracting with non-state entities, including non-government organizations (NGOs), as a way to improve healthcare delivery, and concluded that contracting for delivery of primary care can be very effective and that improvements can be rapid(13)

PBF was intended to contribute to the improvement of health providers’ performance and, ultimately, improve the quality of health service delivery at the operational level. At the same time, it means a fundamental change in the way the health sector is financed, with a shift from input to output funding. This requires changes in accountability structures and the concomitant redistribution of tasks and responsibilities among different actors.(12)

Boosting performance and quality of healthcare delivered to beneficiaries were the main reasons for performance-based financing. The principle of autonomy is central to PBF, whereby providers directly negotiate and finance their performance contracts(8)

Nkambe Health District was one of the districts in the North West Region of Cameroon who piloted PBF in 2012.This study assessed PBF’s effect on obstetric care quality (antenatal, intrapartum, postnatal), provider knowledge/practices, and satisfaction, contributing evidence on the effects of PBF at the subnational (district) level in Cameroon

## 2. Method

### 2.1. Study Design and Setting

We employed a descriptive cross-sectional study from December 2016–July 2017 in Nkambe Health District (North Region, Cameroon).

### 2.2. Participants and Sampling

Simple random sampling of 315 women (who have received prenatal/intrapartum/postnatal care

≤6 weeks before the study) and convenience sampling of 23 providers (≥2 years PBF experience).

### 2.3. Data Collection

Two structured questionnaires (women/providers) were administered by 4 trained collectors. Women: service utilization, satisfaction (Likert).

Providers: knowledge/practices (WHO standards), satisfaction.

Ethical approval: Administrative and ethical clearance was obtained from the North West Regional Delegation of Public Health and Northwest Regional Ethics Committee. Verbal consent was obtained from all participants prior to data collection

### 2.4. Variables

Primary Outcome: Obstetric care quality composite (ANC visits ≥4, partogram use, institutional delivery, PNC attendance).

Exposures: PBF exposure (duration), sociodemographic. Covariates: Proximity, education, providers’ cadre.

### 2.5. Data analysis

Data was analyzed using SPSS v20.Descriptive statistics (means/SD, frequencies), χ^2^ (associations), and Spearman’s correlations. p<0.05 was considered significant. Thematic analysis for open responses. Quality scored per WHO guidelines (e.g., partogram completeness)

## 3. Results

The results obtained are presented below.

### 3.1. Sociodemographic characteristics of participants

Women (n=315)

Mean age 26±5 years; 65.9% married; 49.5% primary education; 36.6% farmers as shown in Table 1

**Table 1.**
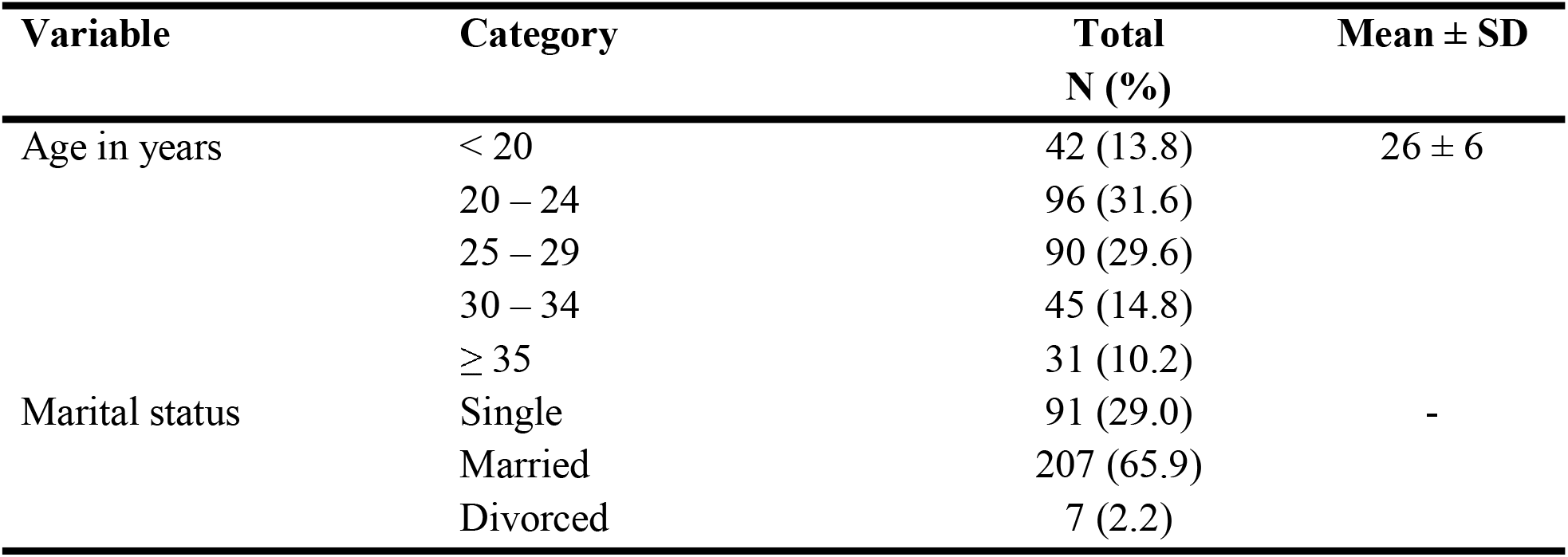
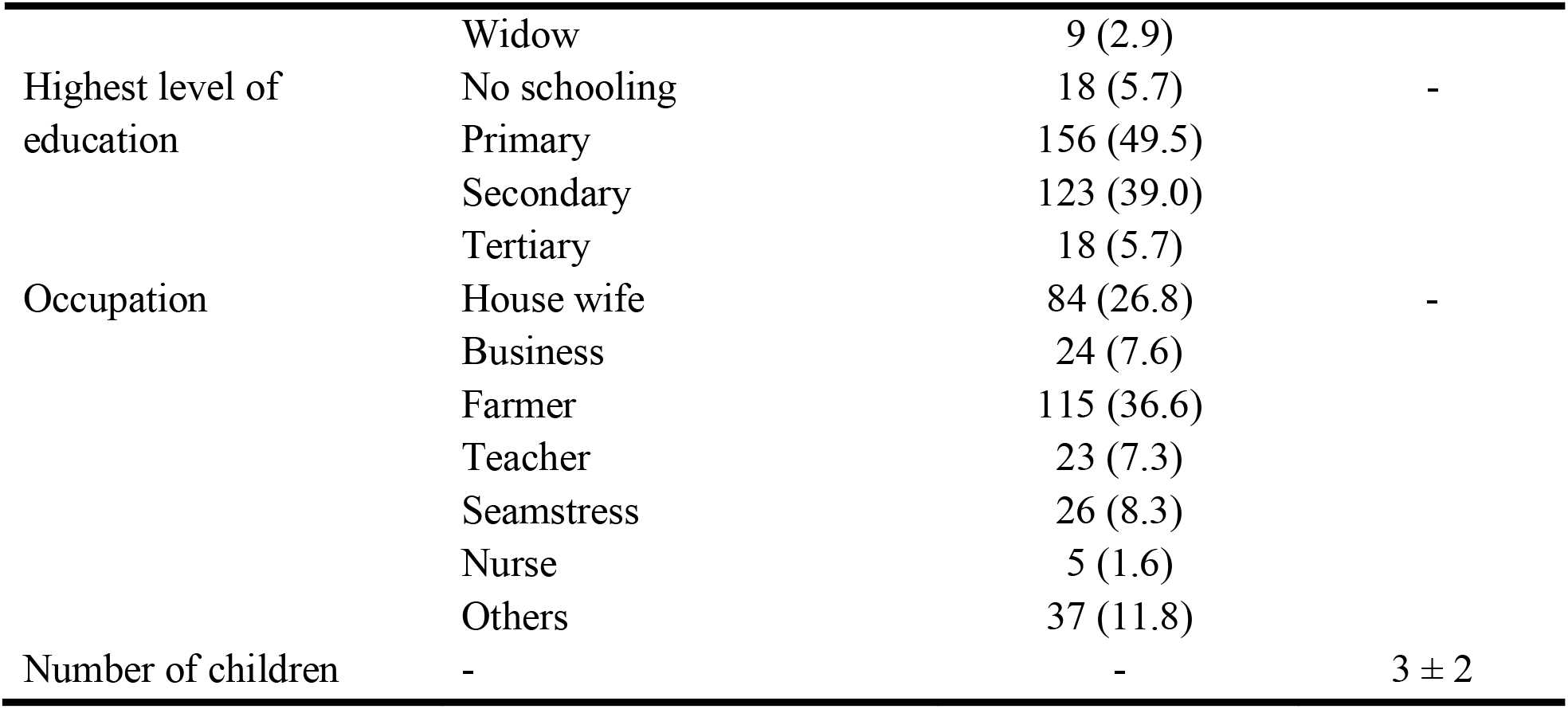
Sociodemographic characteristics of mothers (n = 315)

Service providers (n=23)

59.4% aged 25-34; 68.8% female; 37.5% SRNs; mean experience 7±7 years as shown in Table 7

### 3.2. Service Utilization

93% institutional deliveries (preferred for quality care 84.9%, previous ANC 9.3% and proximity 4.1%); as shown in Table 2.

**Table 2.** Service utilization (n = 315)

| Variable | Category | Total<br>N (%) | Mean ±<br>SD |
| --- | --- | --- | --- |
| Distance from health facility | Minutes trekking | 172 (54.6) |  |
|  | Hours trekking | 58 (18.4) |  |
|  | Motor bike/car | 85 (27.0) |  |
| Birth place of last child | Health facility | 293 (93.0) |  |
|  | Home | 17 (5.4) |  |
|  | Traditional birth attendant | 5 (1.6) |  |
| Reasons for:<br>Health facility | Better quality care | 247 (84.9) |  |
|  | Good reception | 2 (7.0) |  |
|  | Proximity | 12 (4.1) |  |
|  | Attended ANC | 27 (9.3) |  |
|  | Short duration of labor | 1 (0.3) |  |
| Home | Good reception | 2 (100.0) |  |
| Traditional birth attendant | Proximity | 14 (100.0) |  |
| Did you attend ANC before<br>delivery? | Yes | 314 (99.7) |  |
|  | No | 1 (0.3) |  |
| Number of ANC visits | < 4 | 54 (18.0) | 5 ± 1 |
|  | ≥ 4 | 246 (82.0) |  |
| Mode of delivery | NVD | 302 (96.2) |  |
|  | C/S | 12 (3.8) |  |
| Indications for C/S | Macrosomia | 3 (27.3) |  |
|  | Acute fetal distress | 2 (18.2) |  |
|  | Prolonged labor | 3 (27.3) |  |
|  | Malpresentation | 1 (9.1) |  |
|  | CPD | 1 (9.1) |  |
|  | Previous scar | 1 (9.1) |  |
| Number of days spent postpartum | 1 – 2 days | 159 (53.2) | 3 ± 2 |
|  | 3 – 6 days | 124 (41.5) |  |
|  | ≥ 7 days | 16 (5.4) |  |
| Did you revisit the health facility after delivery? | Yes | 291 (93.0) |  |
|  | No | 22 (7.0) |  |
| Reasons for postpartum visits | RDV | 22 (7.5) |  |
|  | Check up | 13 (4.4) |  |
|  | Vaccination | 241 (82.3) |  |
|  | Sick child | 17 (5.8) |  |
| Vaccination status of child | Vaccinated | 307 (97.5) |  |
|  | Not vaccinated | 8 (2.5) |  |
| Reasons for haven't vaccinated child | Child died | 2 (18.2) |  |
|  | Poor sensitization | 4 (36.4) |  |
|  | Vaccines out of stock | 5 (45.5) |  |

99.7% attended at least 5 ANC visits (mean 5±1); 96.2%; Normal vaginal, C-section 3.8% (due to macrosomia (27.3%) and prolonged labor (27.2)).PNC attendance was 93.0 % mostly due to vaccination (82.3%) as shown in Table 3

**Table 3.**
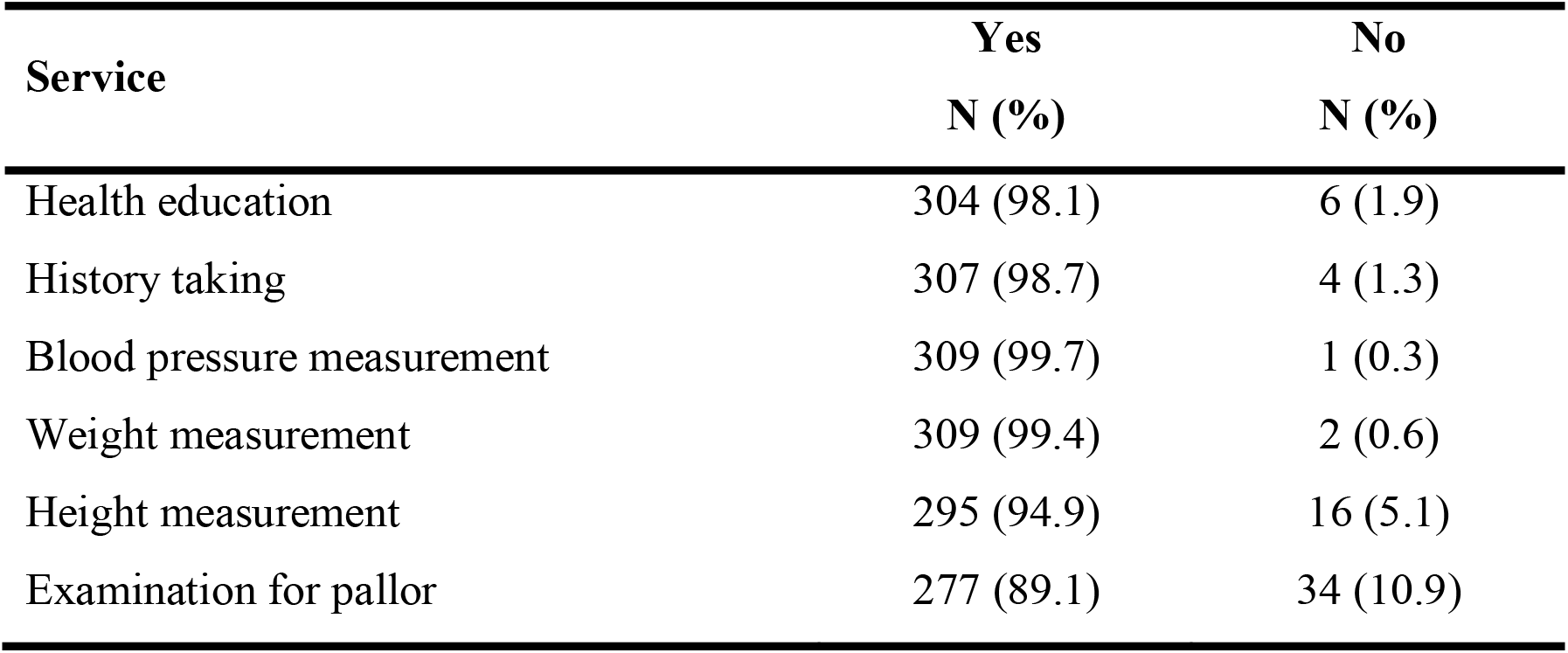
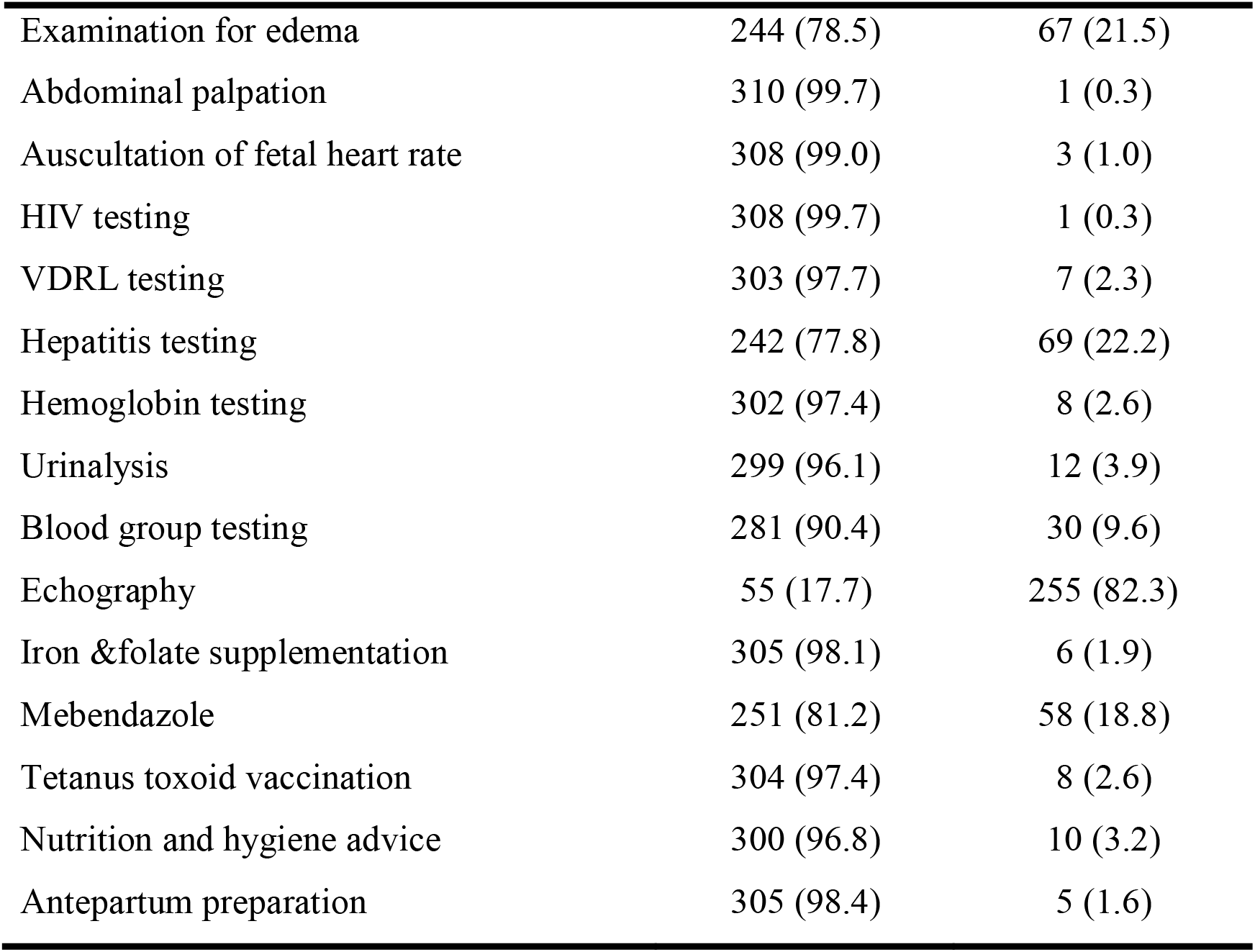
Service offered during ANC (n = 315)

### 3.3. Service Satisfaction and associated factors

#### Consumers

In Table 4, 93.7% reported that ANCs was affordable; 90.8% acceptable quality.

**Table 4.**
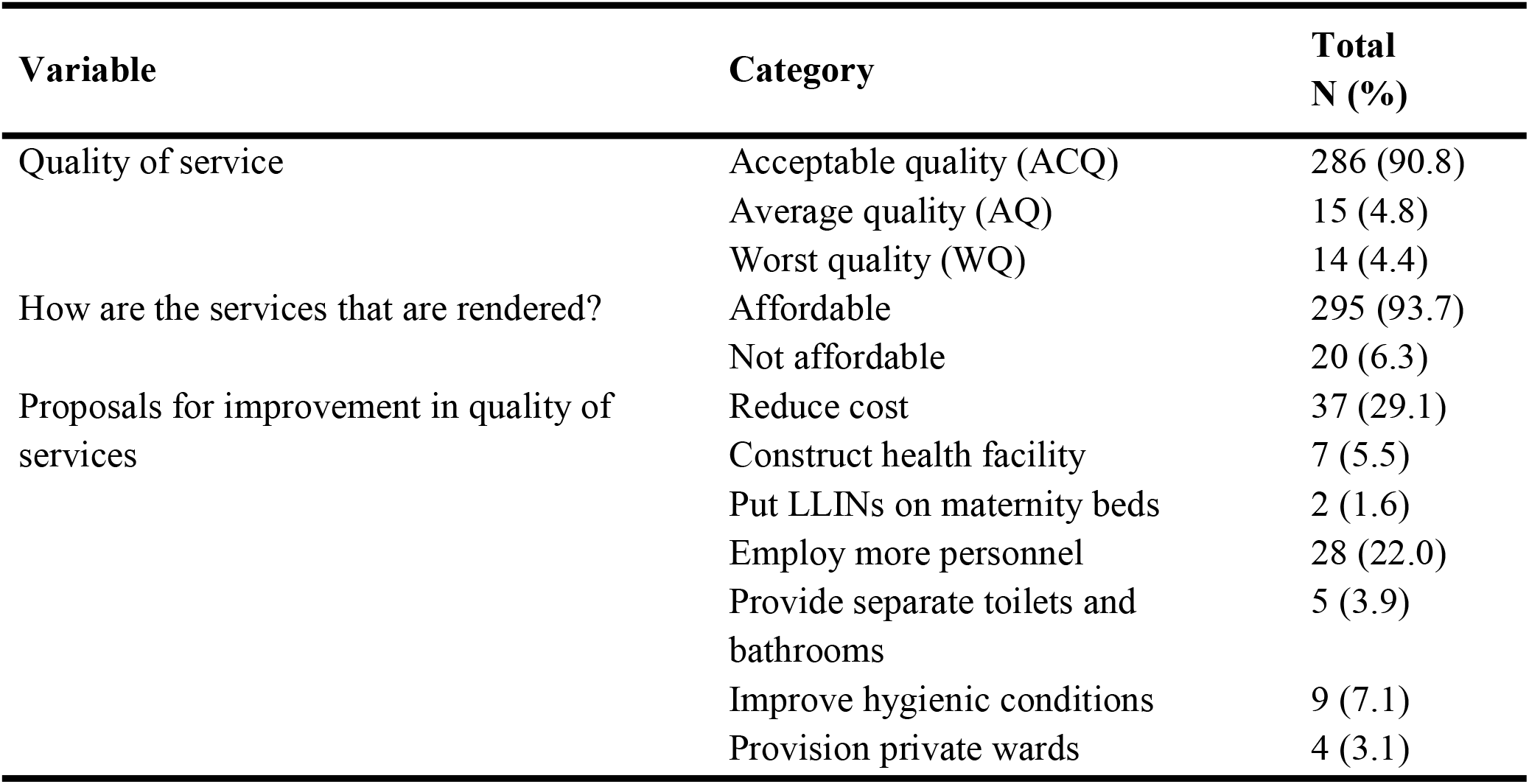
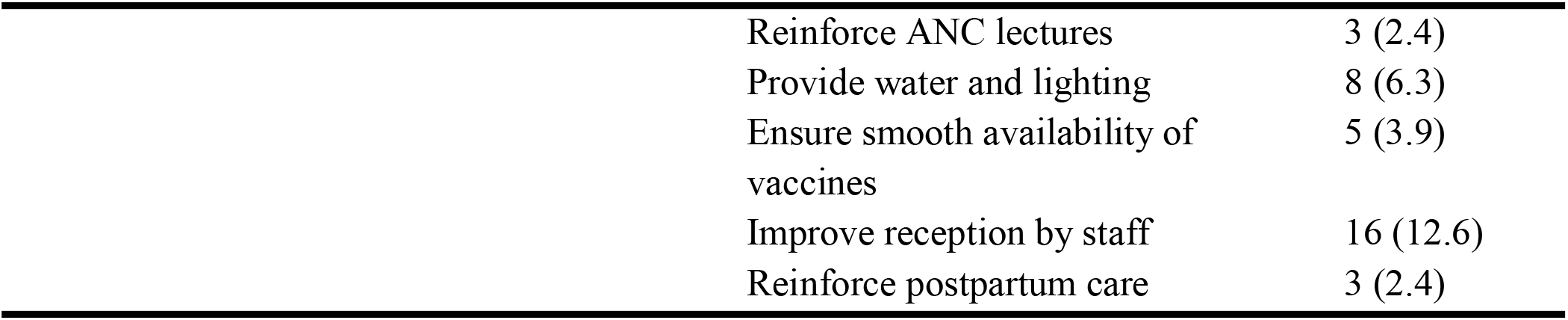
Consumers’ satisfaction of services offered (n = 315)

| Variable | Category | Total<br>N (%) |
| --- | --- | --- |
| Quality of service | Acceptable quality (ACQ) | 286 (90.8) |
|  | Average quality (AQ) | 15 (4.8) |
|  | Worst quality (WQ) | 14 (4.4) |
| How are the services that are rendered? | Affordable | 295 (93.7) |
|  | Not affordable | 20 (6.3) |
| Proposals for improvement in quality of services | Reduce cost | 37 (29.1) |
|  | Construct health facility | 7 (5.5) |
|  | Put LLINs on maternity beds | 2 (1.6) |
|  | Employ more personnel | 28 (22.0) |
|  | Provide separate toilets and bathrooms | 5 (3.9) |
|  | Improve hygienic conditions | 9 (7.1) |
|  | Provision private wards | 4 (3.1) |

|  |  |
| --- | --- |
| Reinforce ANC lectures | 3 (2.4) |
| Provide water and lighting | 8 (6.3) |
| Ensure smooth availability of vaccines | 5 (3.9) |
| Improve reception by staff | 16 (12.6) |
| Reinforce postpartum care | 3 (2.4) |

However, reducing costs (29.1%), increasing staffing strength (22.0%), and improving staff attitudes toward client reception (12.6%) were recommendations to strengthened service quality

#### Service providers

93.8% of service providers were self-satisfied with the obstetric services rendered. 43.7% believed that PBF has greatly improved the quality of obstetric care, while 56.3% also thought that the care had improved, as shown in Table 9.

Proximity to health facility (p=0.017) and attendance of at least four ANCs (p=0.01) were associated with better quality of maternal and child health services received by consumers, as shown in Table 5.

**Table 5.** Relationship between Service quality and other indicators.

| Determinants |  | Service quality |  |  | P-value |
| --- | --- | --- | --- | --- | --- |
|  |  | Acceptable | Average | Worst |  |
|  |  | N (%) | N (%) | N (%) |  |
| Distance from HF | Close | 163 (94.8) | 6 (3.5) | 3 (1.7) | 0.017* |
|  | Far | 123 (86.0) | 9 (6.3) | 11 (7.7) |  |
| ANC attendance | Yes | 283 (91.3) | 13 (4.2) | 14 (4.5) | 0.01* |
|  | No | 3 (60.0) | 2 (40.0) | 0 (0.0) |  |
| Number of ANC's | < 4 ANC's | 49 (17.9) | 4 (30.8) | 1 (7.7) | 0.305 |
|  | ≥ 4 ANC's | 225 (82.1) | 9 (69.2) | 12 (92.3) |  |
| Level of education | No schooling | 16 (88.9) | 2 (11.1) | 0 (0.0) | 0.291 |
|  | Schooling | 270 (90.9) | 13 (4.4) | 14 (4.7) |  |

\*Relationship is significant at level of $p < 0.05$

**Table 6.** Correlation between quality of MCH services and other determinants.

| Determinant | Range | Mean ± SD | Correlation | p-value |
| --- | --- | --- | --- | --- |
|  |  |  | coefficient |  |
|  |  |  | (spearman's rho) |  |
| Number of MCH services rendered | 20 | 18 ± 3 | 0.009 | 0.872 |
| Number of ANC's | 10 | 5 ± 2 | -0.062 | 0.279 |
| Parity (number of | 9 | 3 ± 2 | -0.114* | 0.045 |

children)
\* Correlation is significant at the 0.05 level (2-tailed)

### 3.4. Quality of mother and Newborn service delivery

An assessment of good knowledge and practice in Table 8 showed 96.9% for prenatal services and family planning, and 100% for ANC cards utilization.

**Table 7.** Sociodemographic characteristics of health care providers (n = 32)

| Variable | Category | Males<br>N (%) | Females<br>N (%) | Total<br>N (%) | Mean ±<br>SD | P-value |
| --- | --- | --- | --- | --- | --- | --- |
| Age | 15-24 yrs | 0 (0.0) | 2 (9.1) | 2 (6.2) | 33±9 | 0.593 |
|  | 25-34 yrs | 6 (60.0) | 13 (59.1) | 19(59.4) |  |  |
|  | 35-44 yrs | 4 (40.0) | 7 (31.8) | 11(34.4) |  |  |
| Job title of<br>HCPs | SRN | 3 (30.0) | 9 (40.9) | 12(37.5) |  | 0.096 |
|  | Nursing<br>assistants | 3 (30.0) | 8 (36.4) | 11(34.4) |  |  |
|  | Senior nurses | 1 (100.0) | 3 (13.6) | 4(12.5) |  |  |
|  | Medical<br>doctors | 3 (30.0) | 0 (0) | 3(9.4) |  |  |
|  | midwives | 0 (0%) | 2 (9.1) | 2(6.3) |  |  |
| Years of<br>working<br>experiences | ≤ 3yrs | 5 (50.0) | 12 (54.5) | 17(53.1) | 7±7 | 0.709 |
|  | 3-6 yrs | 2 (20.0) | 4 (18.2) | 6(18.8) |  |  |
|  | 7-9 yrs | 0 (0.0) | 2 (9.1) | 2 (6.2) |  |  |
|  | ≥ 10 yrs | 3 (30.0) | 4 (18.2) | 7(21.9) |  |  |

**Table 8.** Quality of mother and newborn service delivery.

| Variable | Category | Total<br>N (%) | Mean ± SD |
| --- | --- | --- | --- |
| Use ANC cards | Yes | 32 (100) |  |
|  | No | 0 (0.0) |  |
| Number of ANC care provided |  |  | 4±1 |
| Knowledge and practice on ANC | Good | 31 (96.9) |  |
|  | Poor | 1 (3.1) |  |
| Family planning initiation | Satisfactory | 31 (96.1) |  |
|  | Unsatisfactory | 1 (3.1) |  |
| Barriers in family planning | Poor access | 12(37.5) |  |
|  | Husband refusal | 10 (31.3) |  |
|  | Religious practices | 8 (25.0) |  |
| Proportion of women who choose a method |  |  | 4±2 |
| Proportion of MCH staff | Qualified | 31(96.7) |  |
|  | Unqualified | 1(3.1) |  |
| Labour and delivery care | partograph | 28(87.5) |  |
|  | Others | 4(12.5) |  |
| Knowledge and practice | Good | 32(100) |  |
| Postpartum care | Good | 32(100) |  |
| Quality of MCH care with PBF | Markedly improved | 14 (43.8) |  |
|  | Improved | 18 (56.3) |  |
| Personal satisfaction of service rendered by HCP | Yes | 30 (93.8) |  |
|  | No | 2 (6.3) |  |

**Table 9.**
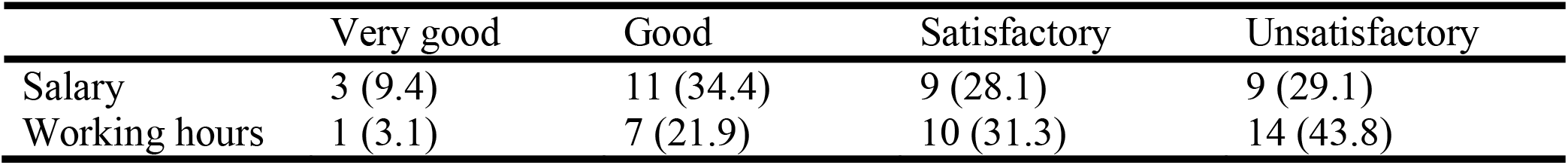
Level of job satisfaction by HCPs.

|  | Very good | Good | Satisfactory | Unsatisfactory |
| --- | --- | --- | --- | --- |
| Salary | 3 (9.4) | 11 (34.4) | 9 (28.1) | 9 (29.1) |
| Working hours | 1 (3.1) | 7 (21.9) | 10 (31.3) | 14 (43.8) |

Postpartum Haemorrhage (PPH) (25%), abortion (15.6%), and malaria (n=4, 12.5%), were the common causes of maternal deaths reported. Late referral (34.4%) and late arrival (n=18, 56.3%) to the referral health facility were the main contributory factors were

Quality score was 100% for intra-partum care;partogram utilization was 87.5%.Postpartum care was 100% (Table 8)

## 4. Discussions

This study shows that Performance-Based Financing has improved the quality of obstetric service delivery in the Nkambe health district. The assessment of knowledge and practice among maternity staff on new WHO guidelines and focused ANC was 97%, consistent with a study on determinants of quality by adeyi(6,14–16). This finding was further reinforced by the mothers’ survey, which showed that 99.7% of pregnant women had at least 5 prenatal visits before delivery(17–20). Moreover, a positive correlation was observed between the quality of services rendered and number of prenatal visits. Attendance of at least four ANCs and schooling were associated with better quality of obstetric care, as demonstrated by Ekenja,Mbah, and others(21– 25). Women living closer to the health facility experienced better service quality than those living farther, with a significant relationship. These results were higher than those of studies conducted in Nigeria, Uganda, and Kenya, which showed lower knowledge and practice among obstetric care providers regarding prenatal and intra-partum care. This difference may be due to the actual established quality of prenatal service delivery, influenced by the PBF scheme compared to other settings. The study also showed that 98% mothers received health education during ANC consistent with a similar study carried out in Maryland (90%) by (Kamila et al., 2007).

In addition, 99.7% mothers had their blood pressure and weight measured during prenatal visits, which was higher than the results of the 2005 Ethiopian demographic health survey conducted in 2005 (59.2% and 43.7%) and also in another study in North West Ethiopia by Tadese et al., in 2013, which were 8.9% and 4.6%, respectively. The difference may be due to PBF on quality-of-service delivery or differences in the study settings.Meanwhile 89% were examined for anemia compared to 61.2% were tested for hemoglobin in a similar study in Ethiopia by Tadese et al. in 2013.Abdominal palpation and fetal heart auscultation were performed in the majority of pregnant women (99.7% and 99.0%,) respectively, consistent with a similar study conducted in Ethiopia by Tadese et al. in 2013. Furthermore, 99.7% were screened for HIV/AIDS, 79% for hepatitis B, (17.7%) did at least one abdominal ultrasound before delivery.

A similar study in Tanzania showed that some examinations were performed very regularly (weighing, auscultation of the fetal heartbeat, and palpation of the fundus) for a greater proportion of the clients (99%).

Performance-based financing has markedly improved the quality of intra partum service delivery. The proportion of qualified staff in maternity was 97%, with the majority having the state enrolled nursing diplomas. The rate of implementation of labor practices like active management of the third stage of labor, was 100%, while 88% utilized the partogram to monitor labor. The results were higher those of the study carried out by Neama et al. in 2012 (49.7%) on the level of education of maternity staff and years of work experience. These results were also consistent with a similar study by Gurung 2008 and another study in Bagdad (18),which showed that the quality of newborn care is related to nurses’ level of education. The researcher believes that the results are better because of the impact of PBF in these health facilities. The results may also be different because of differences in the study settings or the experience and methodology of the researchers.

Furthermore, the proportion of maternity staff with adequate knowledge and practice of post-natal care was 100% backed by a similar study in England that training impacts knowledge and confidence(20) but higher than a similar study carried out in Malawi by Lydia et al. on the quality of post-natal service delivery and supported by 93% women re-visits to the health facility after delivery for child routine vaccination (98%), consultations, medical checkups, family planning, or for an appointments.

Mother and newborn service utilization has increased with the advent of PBF. The study revealed that most women (99%) underwent ANC before delivery. In addition, the rate of institutional delivery was 93%, whereas postpartum visits was 93%. Better quality care and proximity to the health facilities were the main reasons advanced for service utilization, contrary to findings from 2013 Nigeria Demographic and Health Survey (DHS), wherein at least four ANC visits were 54% and 37%, delivery(26). Maternal education was related to service utilization, although this relationship was not strongly established in this study, in contrast with a systematic review on drivers and deterrents, which showed that maternal education and awareness of pregnancy risk factors promote facility delivery in sub-Saharan Africa(27), Ethiopia(25), and Cameroon(23). A study in Ethiopia also found that institutional delivery was associated with the attendance of at least two ANC and maternal knowledge. These results are consistent with a similar study carried out in Ethiopia(22) (94%).

This study also showed that the quality of care had a significant relationship with proximity to the health facilities (94.8%). These findings are similar to those of a study in rural Ghana, which found that contraceptive use and facility delivery were less likely among women living further than 2 km and 1 km, respectively, from a facility compared to women living within 2 km and 1 km of a facility(28,29). Also the level of mothers’ satisfaction was 96%, while 94% could afford the cost of essential services rendered. These results were higher than similar those carried out by Rahel Tesfaye et al. in 2016 and 2017 in Ethiopia (79.1% and 52.6%), West India (86%), and Wolaita (82.9%).These variations in results may be attributed to actual differences in the quality of service delivery, which is the effect of performance-based financing. This could also be due to differences in the study settings.

However, cost reduction (29%), recruitment of more personnel (22%), and improvement of attitudes by staff toward clients were recommendations for service improvement.

The study also revealed that the majority (71%) of health workers were satisfied with their pay package, workload was slightly reduced, and the majority of health staff fully understood their job description, similar to findings made by Manonji, nguileifem(30–32).

## 5. Conclusion

Performance-based financing (PBF) has demonstrated significant improvements in the quality of maternal and child health services in Nkambe health district in the north west region of Cameroon Key study findings indicate enhanced prenatal care, with 99.7% of pregnant women receiving at least 5 antenatal visits. Intra-partum care quality also improved, with 97% of deliveries conducted by qualified personnel and 100% implementation of labor room best practices. Post-natal care showed similar improvements, with 100% of the staff concerned demonstrating adequate knowledge and practice. Service utilization also increased markedly, with 99% ANC attendance, 93% institutional deliveries, as well as 93% postpartum visits.

consumer satisfaction rate was 96%, while 94% found services affordable. The PBF scheme also positively impacted health workers’ motivation, with 71% reporting satisfaction with their pay packages.

However, some areas still require attention, including cost reduction, staff recruitment, and positive staff attitudes toward clients.

This study recommends scaling up the PBF scheme nationwide and potentially adopting it as a national health financing policy. Further research is needed to explore PBF’s impact of PBF on specific aspects of obstetric care in different regions.

In conclusion, PBF has proven to be an effective strategy for improving maternal and child health services in the Nkambe Health District, addressing key challenges in service quality, utilization, and health worker motivation. Its continued implementation and expansion could significantly contribute to the realization of MDGs four and five in Cameroon.

## Ethical approval and informed consent

This study received ethical clearance from the Regional Ethics Committee for Human Health (Reference No: 750/CERSH/2016) and administrative authorization from the Regional Delegation of Public Health, North Region, Cameroon. The study protocol, data collection instruments, and participant information sheets were reviewed and approved prior to commencement on December 1, 2016

## Data Access Statement

Research data supporting this publication are available upon request using the

## Author Contributions

EEA conceptualized and designed the study, collected and analyzed data, and drafted the manuscript. EEA, OR contributed to the analysis of the results and to the writing of the manuscript. NVC contributed to the design and implementation of the research. BF supervised the project.

## Declaration of AI and AI-assisted technologies in the writing process

During the preparation of the manuscript, the authors used ChatGPT to improve language and coherence, and the flow of the ideas of the literature. This was followed by reviewing and editing the contents as needed, and taking full responsibility for the content of the published article

